# Epilepsy Classification using the International League Against Epilepsy (ILAE) 2017 classification, in resource limited setting

**DOI:** 10.1101/2022.10.14.22281100

**Authors:** Hajatmena A M Alkhedir, Abubaker A MohamedSharif, Isra Bdraldein Salih Mohammed, Inaam N Mohamed

## Abstract

**Objectives:** The main aim of the study is to classify patients with epilepsy using ILAE classification 2017, and to test whether it can like Sudan.

**Methods:** This is a descriptive, prospective cross sectional study, conducted in two main pediatric epilepsy clinics in Khartoum State-Sudan.

**Results:** This study included 350 patients with a mean age of (8.4 ±4.7) years. Male to female ratio was 1.5:1. The mean age at the onset of seizure was (3.73 ±3.73) years. The mean duration of the illness was (4.71 ±3.91) years. ILAE Classification was applied and 251 (71.11%) had generalized onset seizures, 95 (27.7%) had focal onset and four (1.1%) had unknown seizure onset.

**Significance:** ILAE 2017 is the last updated classification for epilepsy and very few studies were done especially in resource-limited settings.

## Background

The classification of seizures that has been revised and put in modified form since 1981 was encouraged by a lot of factors. Some seizure types, for example tonic seizures or epileptic spasms, can have either a focal or a generalized onset. There are some of the terms that have been used in classification of seizures which are considered socially and publicly unacceptable, e.g. “simple partial,” and “complex partial.”, and some types of seizures are not encompassed in the 1981 classification. The new classification (by ILAE 2017) main new concept is to describe the onset of seizure, awareness in focal seizure, motor versus non-motor onset in focal and generalized seizures. The main aim of this study is to classify patients with epilepsy using ILAE classification 2017, and to test whether it can be applied in resource-limited countries like Sudan.

## Methods

This is a descriptive, prospective cross sectional study, conducted in two main pediatric epilepsy clinics in Khartoum State-Sudan. One at Jafar Ibnauf specialized hospital for children and another at Soba University Teaching Hospital. The Neurology outpatient departments (OPD) clinics are carried out once per week, with a total number of about (30-50) patients per clinic, with patients referred from different states of Sudan. Two pediatric neurologists, three specialists and 4-6 pediatric residents cover the clinic. Pediatric neurologist made the final classification. Study included all patients (age 2 month to 18 years) attending to the OPD in the period from January to April 2020.

### Case definition

Epilepsy: A diagnosis of epilepsy in this study is retained if the patient had at least two or more epileptic seizures unprovoked by any immediate cause [1].

### Epilepsy classification

For each patient, epilepsy has been classified using the ILAE classification 2017. There are factors that are considered in the classification including seizure onset (Focal/Generalized/ Unknown) based on description of signs and sign-using behavior and personal interview; however, we utilized EEG findings and supportive information like recorded video of the event, were also utilized. Patients with focal seizures were also classified into impaired/unimpaired level of consciousness and focal to generalized tonic clonic type. Similarly, focal and generalized onset seizures were classified into motor or non -motor onset. Seizures of unknown onset are considered when tonic–clonic seizures start to be obscured. The seizure was considered unclassified, if the information was inadequate or unable to fit the seizure in any categories.

At the beginning of the study, all patients did an EEG. The EEGs were done using the 10–20 system with photic stimulation and hyperventilation procedures done when indicated; none of the patients had undergone a video-telemetry EEG or an ambulatory EEG recording because these types of EEG are unavailable in the study setting. The EEGs were interpreted and reported by an adult neurophysiologist with special interest in pediatrics EEGs.

Ethical approval: Ethical approval was obtained from Sudan Medical Specialization Board (SMSB) Ethics and Research Committee. Informed written consents obtained from the parents or caregivers after explaining the aim of the study in simple Arabic language.

## Results

This study included 350 patients with a mean age of (8.4 ±4.7) years. The mean duration of the illness was (4.71 ±3.91) years. Male to female ratio was 1.5:1. The mean age at the onset of seizure was (3.73 ±3.73) years.

ILAE Classification was applied and 251 (71.11%) had generalized onset seizures, 95 (27.7%) had focal onset and four (1.1%) had unknown seizure onset. Fifty-three (56.4%) patients with focal onset seizures had intact aware, and forty-one (43.6%) had impaired levels of consciousness. Sixty-seven (70.5%) patients had focal motor onset seizure, 15 (15.8%) non-motor and 13 (13.7%) had focal to bilateral tonic clonic seizures. The focal motor and non-motor seizures were further classified as in table 1 and 2.

**Table 1:** Focal onset seizure classification

|  |  | Type | Number | % |
| --- | --- | --- | --- | --- |
| Focal onset | Aware | Motor | 31 | 58.5 |
|  |  | Non- motor | 14 | 26.4 |
|  |  | Focal to bilateral tonic clonic | 08 | 15.1 |
|  |  | Total | 53 | 55.8 |
|  | Impaired | Motor | 36 | 85.7 |
|  |  | Non- motor | 01 | 02.4 |
|  |  | Focal to bilateral tonic clonic | 05 | 11.9 |
|  |  | Total | 42 | 44.2 |

**Table 2:** Focal onset seizure classification (n=95)

|  |  | Number | % |
| --- | --- | --- | --- |
| Focal | Motor | 67 | 70.5 |
|  | Non motor | 15 | 15.8 |
|  | Focal to bilateral tonic_clonic | 13 | 13.7 |
| Motor | Automatism | 00 | 00.0 |
|  | Atonic | 01 | 01.5 |
|  | Clonic | 47 | 70.1 |
|  | epileptic spasm | 00 | 00.0 |
|  | Hyperkinetic | 00 | 00.0 |
|  | Myoclonic | 14 | 20.9 |
|  | Tonic | 05 | 07.5 |
| Non motor | Autonomic | 01 | 06.7 |
|  | behavior arrest | 01 | 06.7 |
|  | Cognitive | 00 | 00.0 |
|  | Emotional | 06 | 40.0 |
|  | Sensory | 07 | 46.7 |

The generalized onset seizures, 239 (95.2%) were of motor onset and the 12 (4.8%) were of non-motor onset. The generalized motor onset seizures, 106 (44.2%) had tonic clonic seizures, 47(19.6%) tonic, 37(15.4%) clonic, 19 (7.9%) myoclonic, 23 (9.6%) atonic, 7 (2.9%) epileptic spasm and one (0.4%) patients had myoclonic tonic clonic seizures. The generalized non-motor type, eight (72.7%) had atypical seizures and one (9.1%) had typical seizures. Two (18.2%) patients had eyelid Myoclonia. Table 3

**Table 3:** Generalized onset seizure classifications (n=251)

|  |  | Number | % |
| --- | --- | --- | --- |
| Generalized onset | Motor | 239 | 95.2 |
|  | Non motor | 012 | 04.8 |
| Motor | Tonic_clonic | 106 | 44.2 |
|  | Clonic | 037 | 15.4 |
|  | Tonic | 047 | 19.6 |
|  | Myoclonic | 019 | 07.9 |
|  | Myoclonic _tonic _clonic | 001 | 00.4 |
|  | Myoclonic _atonic | 000 | 00.0 |
|  | Atonic | 023 | 09.6 |
|  | Epileptic spasm | 007 | 02.9 |
| Non motor | Typical | 001 | 09.1 |
|  | Atypical | 008 | 72.7 |
|  | Eyelid Myoclonia | 002 | 18.2 |

Three (27.3%) of patients had unknown onset, all presented with tonic clonic convulsions. One patient (9.1%) was unclassified.

## Discussion

ILAE Classification 2017 was applied to patients under this study in which (99.7%) of patients were fully classified. There were few studies done using ILEA classification 2017, so we use this classification in our study.

In this study the majority of patients had generalized onset seizures, one third had focal onset. Badrelddin et al from Sudan2 and Selina H Banu et al from Bangladesh reported similar to this study3 however, they applied ILAE Classification 2010. In contrast to Suvasini et al from India who used ILAE Classification 2017 observed that most common types of seizures were focal onset 5, however he reported similar to this study focal with motor onset type is the commonest.

In this study, most of the patients had generalized motor onset seizure. Suvasini et al from India reported a similar finding4.

Most patients with unknown onset presented with a motor pattern and almost all of them presented in the form of a tonic clonic type similar to the study Suvasini et al and Gowda et al 4, 5.

When epilepsy is diagnosed, Then we come to epilepsy syndrome, where we can make a diagnosis of a specific epilepsy syndrome. ILEA classification 2017 listes the causes with each phase, accentuating the importance of considering the etiology of epilepsy when making the diagnosis, as it conveys deemed treatment results. The etiology of epilepsy has been classified into six subgroups, chosen for their likelihood of therapeutic ramifications. Also in this new classification, new wording is antedated such as epileptic and developmental encephalopathy.

## Data Availability

All data produced in the present study are available upon reasonable request to the authors

## Data Availability statement

The datasets used and/or analyzed during the current study are available from the corresponding author on reasonable request.

## Ethical publication statement

Consent of publication was collected from all the participants in this study as part of the verbal consent.

## Patient consent statement

To ensure adherence to ethical guidelines, several measures were adopted while conducting this study:

1. No incentives were offered to the participants in return for their participation.
2. Verbal consent was obtained from the participants before filling the questionnaire.
3. Participants were informed that their participation in this study is voluntary, no incentives or compensations will be offered in return, and that they have the right to withdraw from the study at any stage. The scientific value of their participation was explained in the verbal consent.
4. The contact information of the principal investigators was provided for participants.
5. All the participants’ information was kept private by keeping it in a secured folder in a password-protected computer owned by the study investigators. No information was shared with any other individuals or entities.

## Authors’ contributions

Hajatmena Alkhedir: conception of this study. Hajatmena Alkhedir designed the study and formulated its methodology. Abubaker Mohamedsharif and Isra Mohammed analyzed the data. Abubaker Mohamedsharif and Isra Mohammed interpreted the data. Abubaker Mohamedsharif contributed to the results section. Isra Mohammed and Inaam Mohamed contributed to the discussion section. Inaam Mohamed drafted the work and substantially revised it. Inaam Mohamed critically reviewed and approved the manuscript.

All authors listed have agreed to both be personally accountable for the author’s own contributions and to ensure that questions related to the accuracy or integrity of any part of the work, even ones in which the author was not personally involved, are appropriately investigated, resolved, and the resolution documented in the literature.

All authors listed read and approved the final manuscript.

